# N-of-1 analysis of circadian data reveals potential for precision chrono-medicine approach of concomitant exercise and metformin recommendations

**DOI:** 10.1101/2024.11.13.24317280

**Authors:** Katy Lyons, Brenda Peña Carrillo, Lara Talia Dasar, Odiaka Philippa Ifeyinwa, Brendan M. Gabriel

## Abstract

The integration of chrono-medicine into disease management has potential for cost-effective improvements, particularly in type 2 diabetes care. While both exercise and metformin are effective in lowering glycaemia, their combined effect is non-additive. Individual circadian rhythms suggest that personalised timing of interventions may optimise outcomes. This study aims to investigate the heterogeneity in response to the timing of exercise and metformin intake using an n-of-1 approach within a randomised crossover trial, thereby exploring the potential for individualised chrono-medicine strategies.

A previously published 16-week randomised crossover study was conducted to explore the potential therapeutic effects of prescribed moderate exercise timings in participants undergoing metformin monotherapy. Physical activity, heart rate, sleep, and glucose levels were tracked using wearable technology and continuous glucose monitors. Data were collected during baseline, and throughout the intervention periods. Analysis focused on individual responses to the timing of exercise and metformin intake.

Morning exercise significantly lowered 24-hour post-exercise blood glucose levels compare to evening exercise. Both exercise timings reduced mean blood glucose levels, but morning exercise had a greater effect (mean difference: -0.63 mmol/L, p<0.001) than evening exercise (mean difference: -0.34 mmol/L, p=0.016).

Individual responses varied, with some participants displaying a substantial reduction in glucose levels in response to morning or evening exercise, while others did not benefit from either exercise intervention. Pre-breakfast metformin intake significantly lowered area under the curve (AUC) glucose values in response to morning exercise compared to post-breakfast, an effect not observed with evening exercise.

Morning exercise combined with pre-breakfast metformin intake is the most effective strategy for lowering blood glucose levels in the greatest number of participants with type 2 diabetes. However, individual response heterogeneity suggests that chrono-medicine approaches must be personalised. Further research is needed to understand the underlying mechanisms of individual variability in response to exercise and medication timing.

**Research in Context:** *What is already known about this subject?:*

- Exercise prescription and metformin treatment are both effective in lowering glycaemia but their combined effect is non-additive.
- It is unknown how individual circadian rhythmicity interacts with these diabetes treatment strategies.

*What is the key question?:*

- Is there potential for personalisation of a chrono-medicine approach to diabetes management?

*What are the new findings?:*

- Morning exercise combined with pre-breakfast metformin intake is the most effective strategy for lowering blood glucose levels in the greatest number of participants with type 2 diabetes.
- Further research is needed to understand the underlying mechanisms of individual variability in response to exercise and medication timing.

*How might this impact on clinical practice in the foreseeable future?:*

- Our study shows that integrating chrono-medicine into disease management has potential for cost-effective improvements, particularly in type 2 diabetes care.

## Introduction

Identifying cost-effective strategies for disease management is crucial. An underexplored area is chrono-medicine, especially pertinent now with wearable devices which can monitor circadian patterns of glucose, heart rate, and physical activity. Growing evidence supports the integration of chrono-medicine into lifestyle and pharmaceutical interventions for disease management (Gabriel & Zierath, 2021). The NHS spends around £10 billion a year on diabetes – around 10% of its entire budget - and it has been suggested that chrono-medicine is a promising approach for cost-effective improvements in type 2 diabetes care (Gabriel & Zierath, 2021).

Exercise and nutritional interventions are well-established as vital and effective methods to manage type 2 diabetes. Several pharmacological treatments are also highly effective and well-tolerated. Metformin, for example, has long been a common intervention and was first used clinically in the 1960s. While metformin is safe and generally well-tolerated, it interacts with lifestyle interventions like exercise, leading to some non-additive outcomes. For instance, both metformin and exercise effectively lower glycaemia when used independently, but the literature suggests that a combinative approach offers no additional improvement (Sharoff *et al*., 2010; Boulé *et al*., 2011; Abdalhk *et al*., 2020). Exercise improves glycaemic regulation partly by enhancing skeletal muscle glucose uptake and has additional benefits such as improving cardiovascular health and reducing cancer risk (Cartee *et al*., 2016). One of the main mechanisms by which exercise promotes beneficial health outcomes is by remodelling skeletal muscle to improve mitochondrial oxidation, for example by increasing total activity of enzymes such as citrate synthase (Leek *et al*., 2001) and therefore improving lipid and glucose oxidation (Gabriel *et al*., 2017; Alhindi *et al*., 2019). However, many of the improved substrate handling effects of exercise appear to be acute, with studies suggesting that some beneficial effects of acute exercise disappear within 48 hours (Gabriel *et al*., 2013). However, many people with type 2 diabetes, including those on metformin, are less likely to exercise regularly (Krug *et al*., 1991). Metformin can reduce exercise capacity and alter perceptions of exercise intensity (Das *et al*., 2018; Kristensen *et al*., 2019), creating a barrier to exercise adherence. Additionally, metformin may interfere with skeletal muscle remodelling after exercise (Konopka *et al*., 2019), potentially hindering beneficial health adaptations. For these reasons, finding ways to encourage exercise adherence and optimise its benefits alongside metformin is essential for improving type 2 diabetes healthcare strategies.

Circadian rhythms are heterogeneous between individuals (Vitaterna *et al*., 2019) and therefore, lifestyle interventions must consider the holistic diurnal environment in which a given individual resides. Thus, when considering the application of precision timing, researchers must consider factors including nutritional intake, physical activity, medicine intake, sleep, and occupation. Researchers have traditionally used observational, cross-sectional, and parallel-group randomised trials to study circadian biology and disease management strategy efficacy. However, these trials may not always outline the most effective treatment for specific subgroups or individual patients. To address the inherent variability in treatment response among clinical populations, n-of-1 trials can be employed (Vieira *et al*., 2017; Samuel *et al*., 2023). These personalised trials contribute to the ultimate aim of all biomedical research, which is to enhance the care provided to individual patients.

Circadian biology presents an excellent area for the application of n-of-1 study designs due to the nature of data collection. The use of wearable technology and repeated time point sampling is a standard procedure across circadian trials and therefore, the statistical design of n-of-1 analysis is appropriate. Therefore, we have used a previously published circadian biology randomised cross-over trial from our laboratory to investigate the heterogeneity of response to our intervention using an n-of-1 approach. We aim to determine the potential for an individual-centred approach when applying chrono-medicine disease management strategies. Our hypothesis is that there are inter-individual glycaemic responses to exercise timing with opposite directionality in people with Type 2 Diabetes being prescribed metformin monotherapy.

## Methods

### Original Study Design

In order to test this hypothesis, we re-analysed data from a previously published study (Carrillo *et al*., 2024). The original study design is described in the previously published paper (Carrillo *et al*., 2024). The study was conducted following ethical approval from the UK Health Research Authority’s Integrated Research Application System (IRAS), and via the London Centre Research Ethics Committee (REC reference: 20/PR/0990; IRAS project ID: 292015) and in accordance with the principles of the Declaration of Helsinki as revised in 2008. Informed written consent was obtained from participants during recruitment. The trial was preregistered – Research Registry Unique Identifying Number: researchregistry6311. Following a randomised, cross-over, two-arm design shown in Figure 1, the trial used Garmin smartwatches and the associated mobile application, Garmin Connect (Garmin Ltd. Olathe, KS, US) to measure physical activity (step count), heart rate and sleep parameters. FreeStyle Libre 2 sensors LibreView software and the LibreLink mobile application were used to record and collect glucose readings in 15-minute intervals. Participants were randomised between exercise intervention groups (morning/evening) Glucose data was collected 2 weeks prior to the start of the first exercise intervention (baseline) and during the first 2 (weeks 1-2) and last 2 (weeks 5-6) of each exercise intervention period. Daily step count, heart rate and sleep were measured throughout the study period. A 4-day food diary was also kept by participants when glucose monitors were worn, and timing and dosage of metformin was recorded. A 2-week wash-out period was included between each intervention period. In the winter cohort of participants, the wash-out period was extended to 4 weeks to avoid crossover into the Christmas holidays. All data sets released for this secondary n-of-1 analysis were de-identified following the principles of GCP (Good Clinical Practice) and GDPR (General Data Protection Regulations).

**Figure 1.**
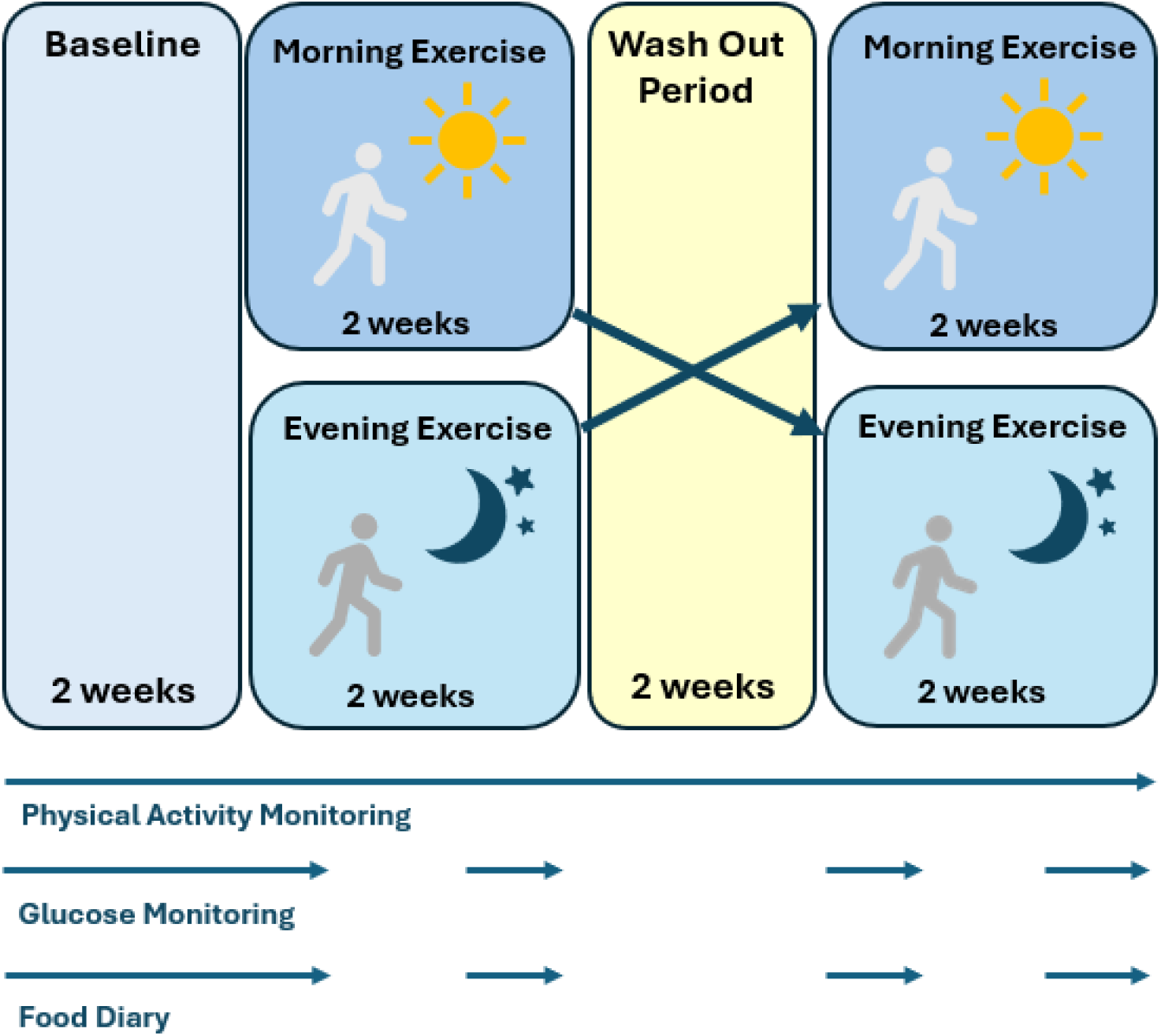
Original Study Design of Trial: n=18 (9 male, 9 female), n=8 morning/evening, n=10 evening/morning. Exercise consisted of 30 mins of moderate intensity walking at 70% max HR every other day. Morning exercise was completed anytime between 7-10am and evening exercise was completed anytime between 4-7pm. Physical activity, heart rate and sleep data were collected throughout the whole study period whereas glucose data was only collected 2 weeks prior (baseline) and the first and last 2 weeks of each intervention (week 1-2 and week 5-6).

### N-of-1 Data Analysis

The analysis of the extracted data incorporates a series of 18 individual n-of-1 analysis which have been used to visualise data in a time-series manner and analyse the optimal timing of metformin intake and exercise routine for each participant (Vieira *et al*., 2017). Participant characteristics, metformin timings, glucose readings, and exercise intervention were extracted from the original randomised cross-over trial. All analysis was carried out on R software 4.3.1 (RStudio 2023.06.2.56) or GraphPad Prism (Version 10.1.12). Exercise intervention periods and control baseline were time matched, setting the first point of the 24-hour day to the start of each exercise period, indicated by the time point ‘0 minutes.’ ‘0 minutes’ corresponds to 7am during the morning exercise intervention and 4pm for the evening exercise intervention.

Time-Series analysis used data from the first 24-hours of each glucose monitoring period (baseline and weeks 1-2 and 5-6 of each exercise intervention) to show each participant’s glucose profile over 24 hours. Missing data was corrected using linear imputation on Microsoft Excel when appropriate. Mean difference meta-analysis was performed firstly on aggregated and averaged data from all participants to analyse the mean difference between daily glucose readings at each 15-minute interval throughout the 24-hours period post-exercise. This analysis was performed to compare each exercise intervention to its time matched baseline period. Secondly, mean difference meta-analysis was used to analyse overall daily mean glucose for each participant, comparing the effect of each exercise intervention and time-matched baseline period. Area Under the Curve (AUC) values were calculated using GraphPad Prism to further compare metformin intake and exercise interventions. Paired student t-tests were performed to test for any statistical significance throughout this study with significance set at p-value<0.05.

## Results

### Summary Characteristics of Study Cohort

A summary of baseline characteristics of the study cohort is shown in Table 1. A total of 18 participants were included in the analysis, with an equal split of male and female participants.

**Table 1.** Baseline Study Cohort Characteristics: Baseline Characteristics were taken from the original study. SEM (Standard Error of the Mean), BMI (Body Mass Index), T2D (type 2 diabetes).

| **Characteristic** | **Mean +/- SEM** |
| --- | --- |
| *Age (years)* | 61 +/- 1.9 |
| *Sex (Male)* | 9 |
| *Sex (Female)* | 9 |
| *BMI (Kg/m<sup>2</sup>)* | 30.7 +/- 0.8 |
| *HbA1c (%)* | 8.0 +/- 0.3 |
| *Time since T2D diagnosis (years)* | 8.2 +/- 1.7 |
| *Time on Metformin (years)* | 5.2 +/- 1.0 |
| *Dose of Metformin (mg)* | 1312.5 +/- 134.6 |
| *Average number of Daily Steps (Baseline)* | 6797.6 +/- 594.6 |

### Time-course of individual glycaemia

Time-series plots show the changes in individual blood glucose levels over the first 24 hours of baseline, morning exercise intervention and evening exercise intervention. 10.06% of all first 24-hour data was missing (Figures 2-3). Of this missing data, 40.75% was appropriately corrected by linear imputation. The remaining 59.25% of missing data was not included in the analysis. Participant 9 and Participant 1 had no data for week 5-6 of evening exercise intervention.

**Figure 2.**
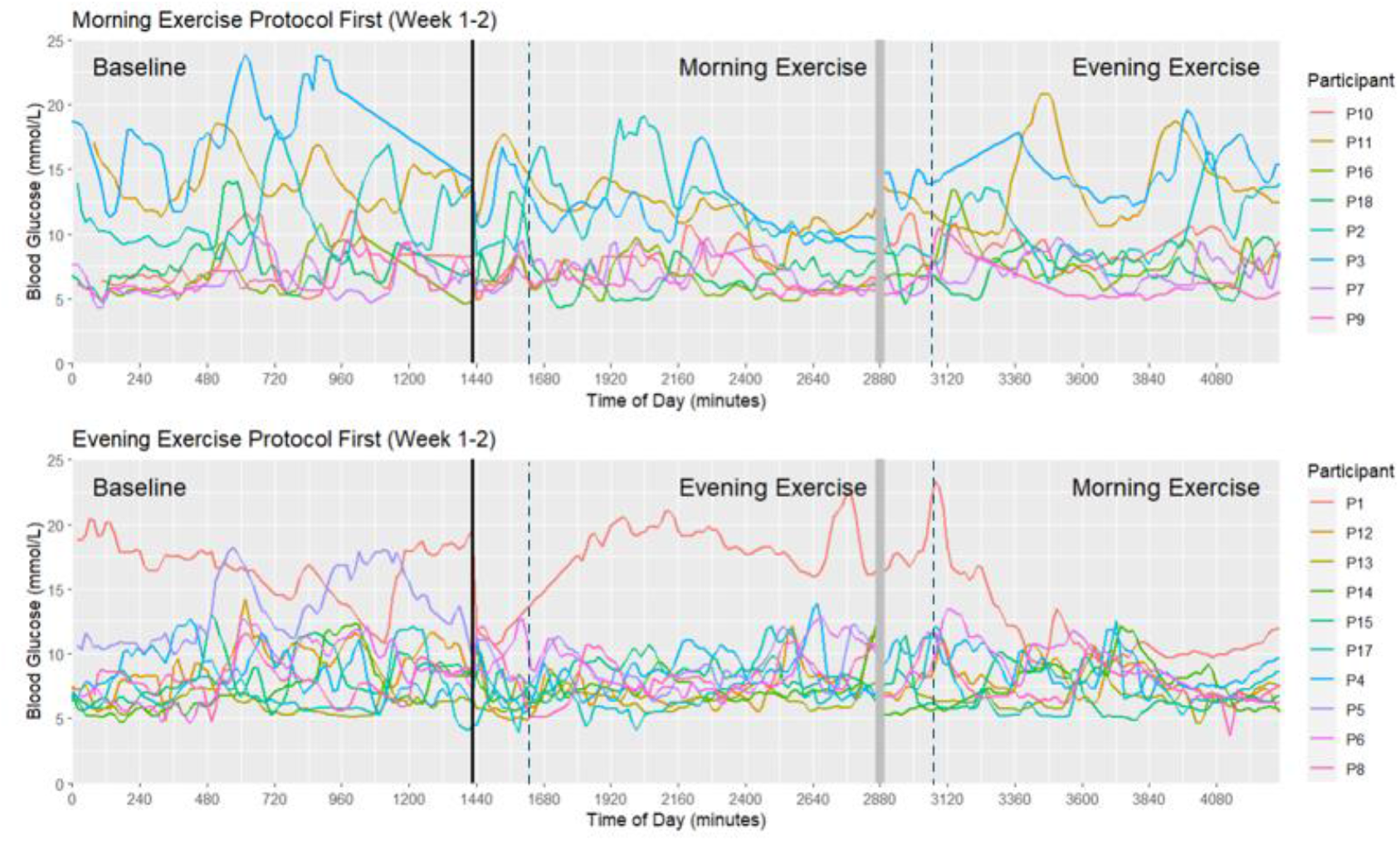
24-Hour Time-Series Plot Showing Week 1-2 of Both Morning and Evening Exercise Intervention: The black line at time point 1440 minutes represents the end of baseline period and grey bar after the first exercise intervention at timepoint 2880 minutes represents the washout period, where no data was collected. Baseline sections follow 24-hour timescale of midnight to midnight, morning exercise from 7am to 7am and evening exercise from 4pm to 4pm. The dash line on plots represents the end of the exercise period. Missing data was corrected by linear imputation where appropriate.

**Figure 3.**
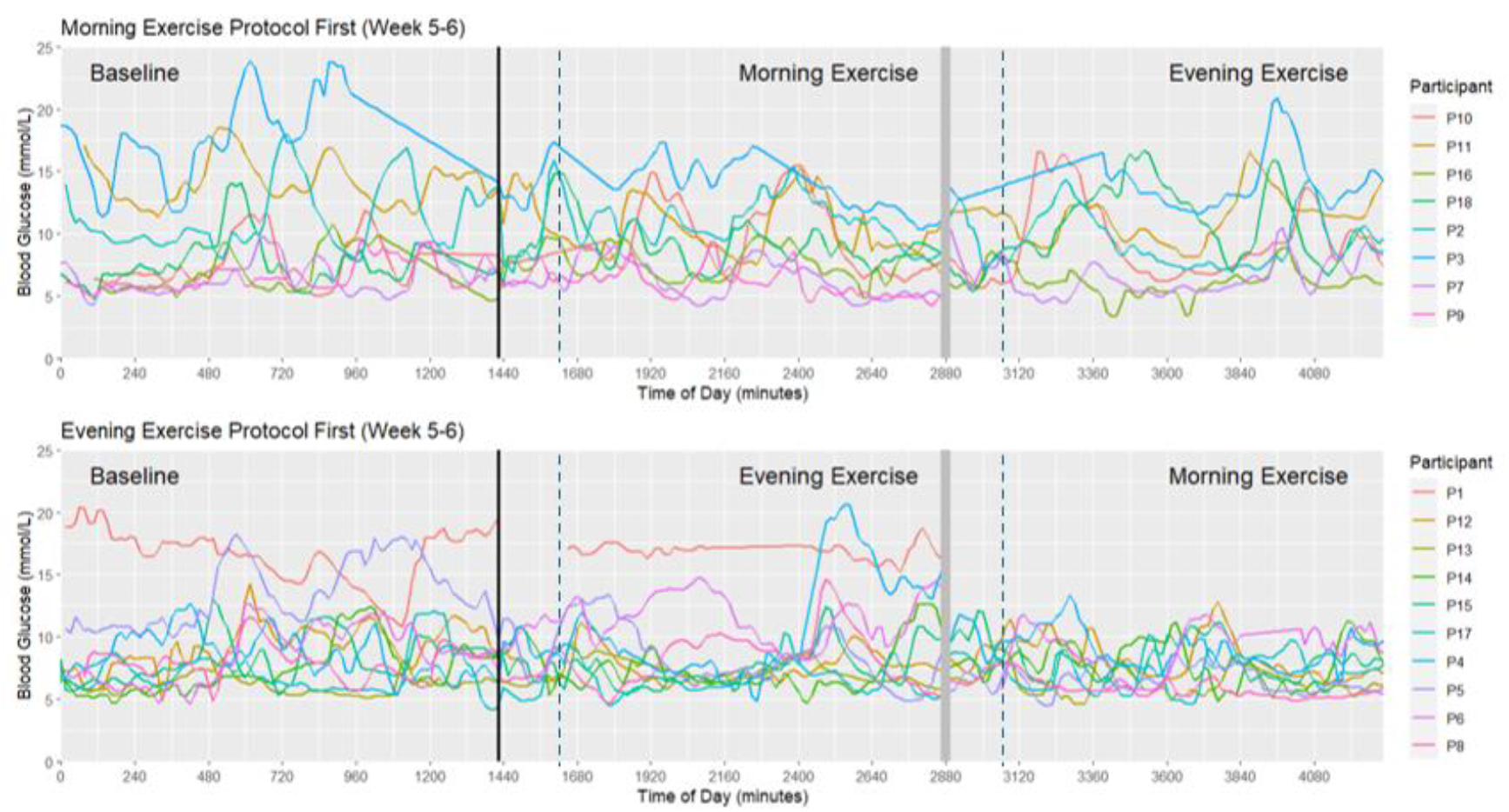
24-hour Time-Series Plot Showing Week 5-6 of Both Morning and Evening Exercise Intervention: The black line at time point 1440 minutes represents the end of baseline period and grey bar after the first exercise intervention at timepoint 2880 minutes represents the washout period, where no data was collected. Baseline sections follow 24-hour timescale of midnight to midnight, morning exercise from 7am to 7am and evening exercise from 4pm to 4pm. The dash line on plots represents the end of the exercise period. Missing data was corrected by linear imputation were appropriate. Participant 9 (P9) had no data present for evening exercise intervention and Participant 1 (P1) was missing data for morning exercise intervention.

### Effect of Exercise on Mean Blood Glucose

Both morning and evening exercise lowered mean blood glucose compared to baseline. Figures 4-5 demonstrate that morning exercise has a greater effect in lowering blood glucose levels over the 24-hour period post-exercise compared to evening exercise. This effect was present over a standard 24-hour period (Figure 4), and also a Zeit-time normalised period (Figure 5), where exercise is classed as Zeit-time 0, 24-hour blood glucose is included from the starting point of exercise. The mean difference meta-analysis presented in figure 6 and figure 7 supports this finding, showing that both morning and evening exercise reduce overall mean levels of blood glucose.

**Figure 4.**
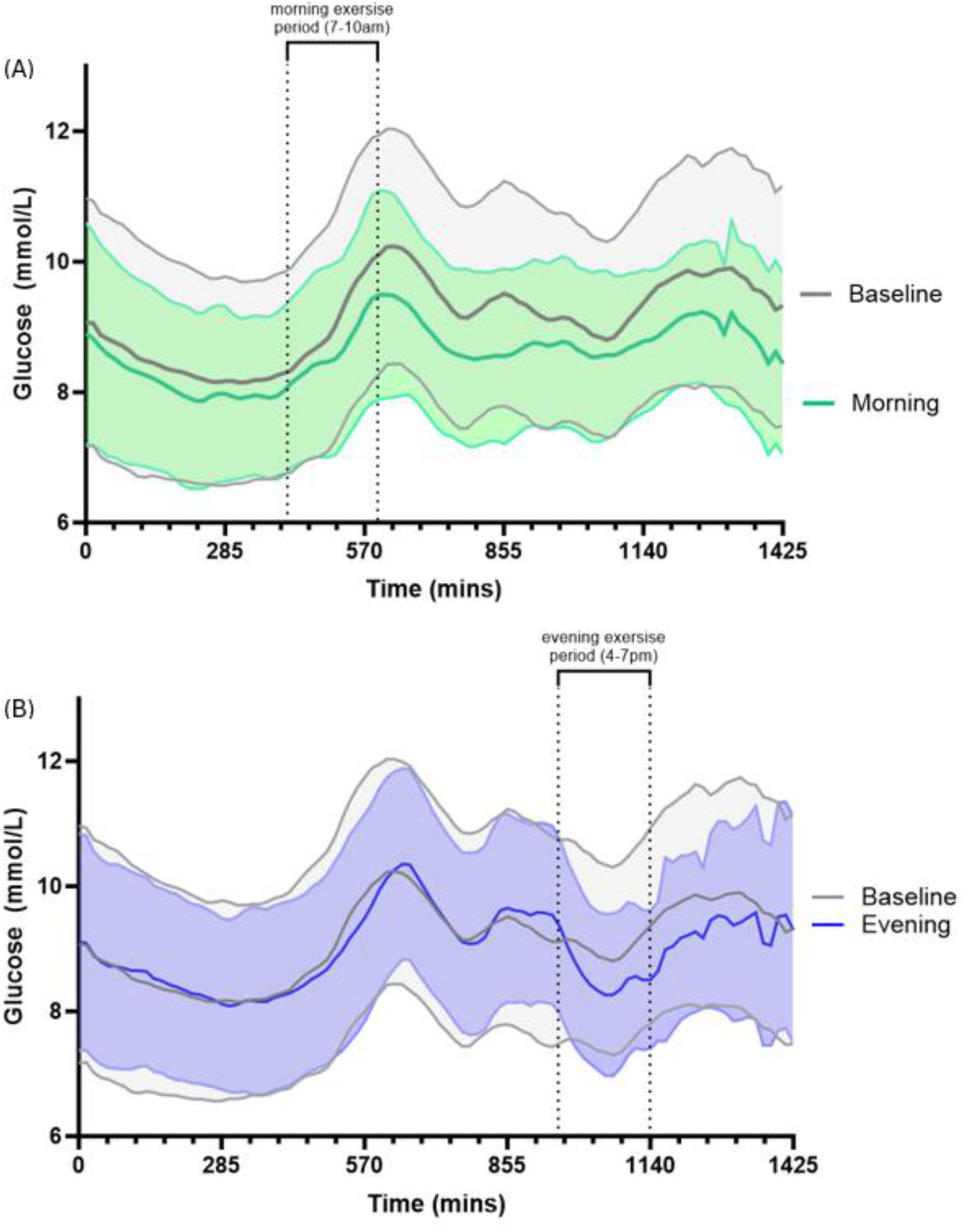
Average Daily Glucose over a 24-hour clock period for each exercise intervention versus baseline: Average blood glucose levels over 24-hour period for each exercise intervention compared to baseline, shown as mean with 95% CI. (A) Morning Exercise Intervention Period: The dash line indicates the window for exercise (7am-10am). (B) Evening Exercise Intervention Period: The dash lines indicate the window for exercise (4pm-7pm). Time 0 mins = 12am and time 1425 mins = 11:45pm.

**Figure 5.**
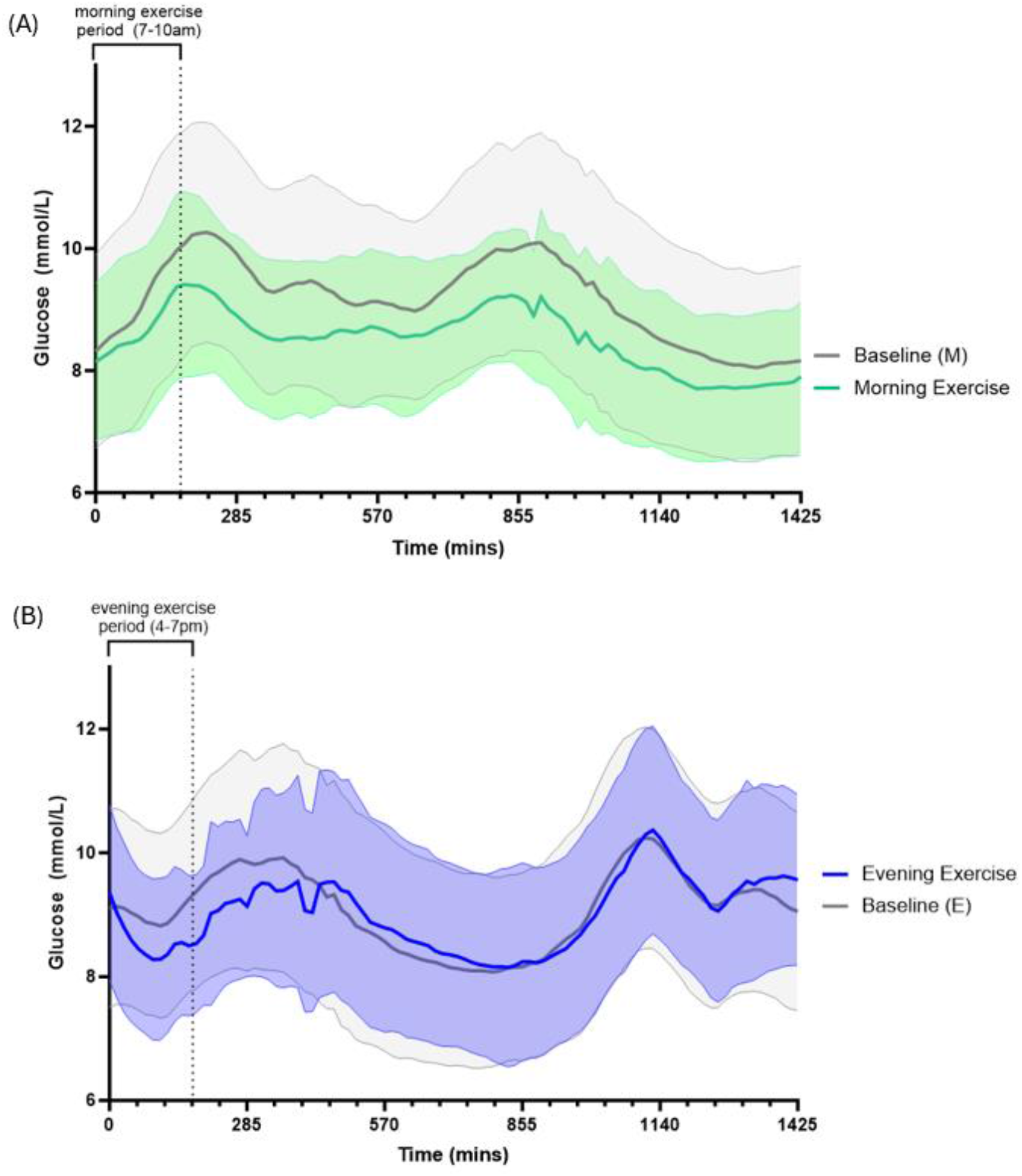
Zeit-time normalised timing of Metformin and the Effect on Individual Average Daily Glycaemic Values. Average blood glucose levels over 24-hour period for each exercise intervention compared to baseline, shown as mean with 95% CI. (A) Morning Exercise Intervention Period: The dash line indicates the window for exercise (7am-10am). (B) Evening Exercise Intervention Period: The dash lines indicate the window for exercise (4pm-7pm).

**Figure 6.**
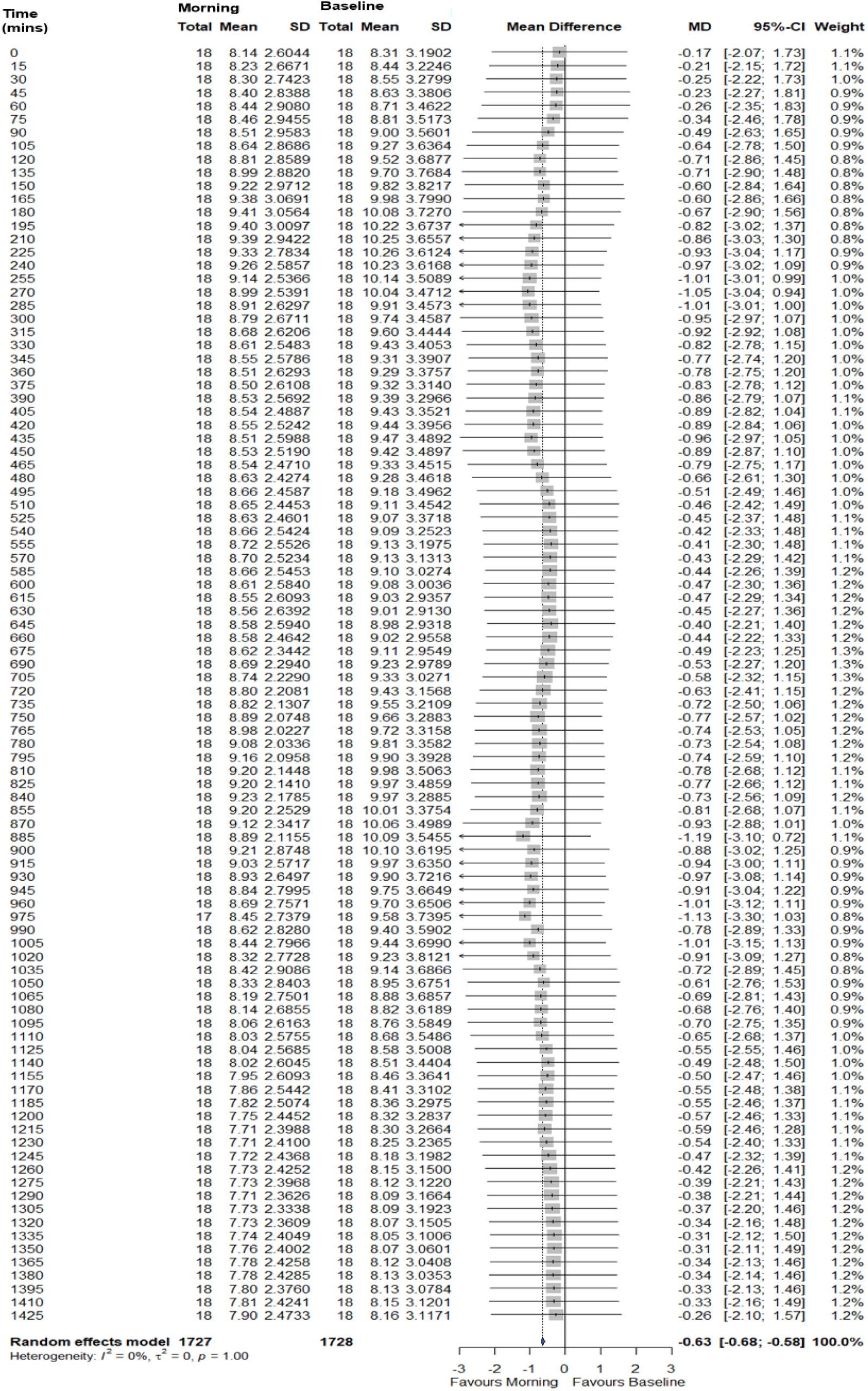
Mean Difference Meta-Analysis Comparing Baseline and Morning Exercise Intervention: 24-hour period is split into 15-minute intervals. Baseline is time matched to morning exercise intervention, therefore 0 mins=7am. Morning exercise period ends at 10am (180 mins). A negative MD favours morning and positive MD favours baseline. SD (standard Deviation), MD (Mean Difference).

**Figure 7.**
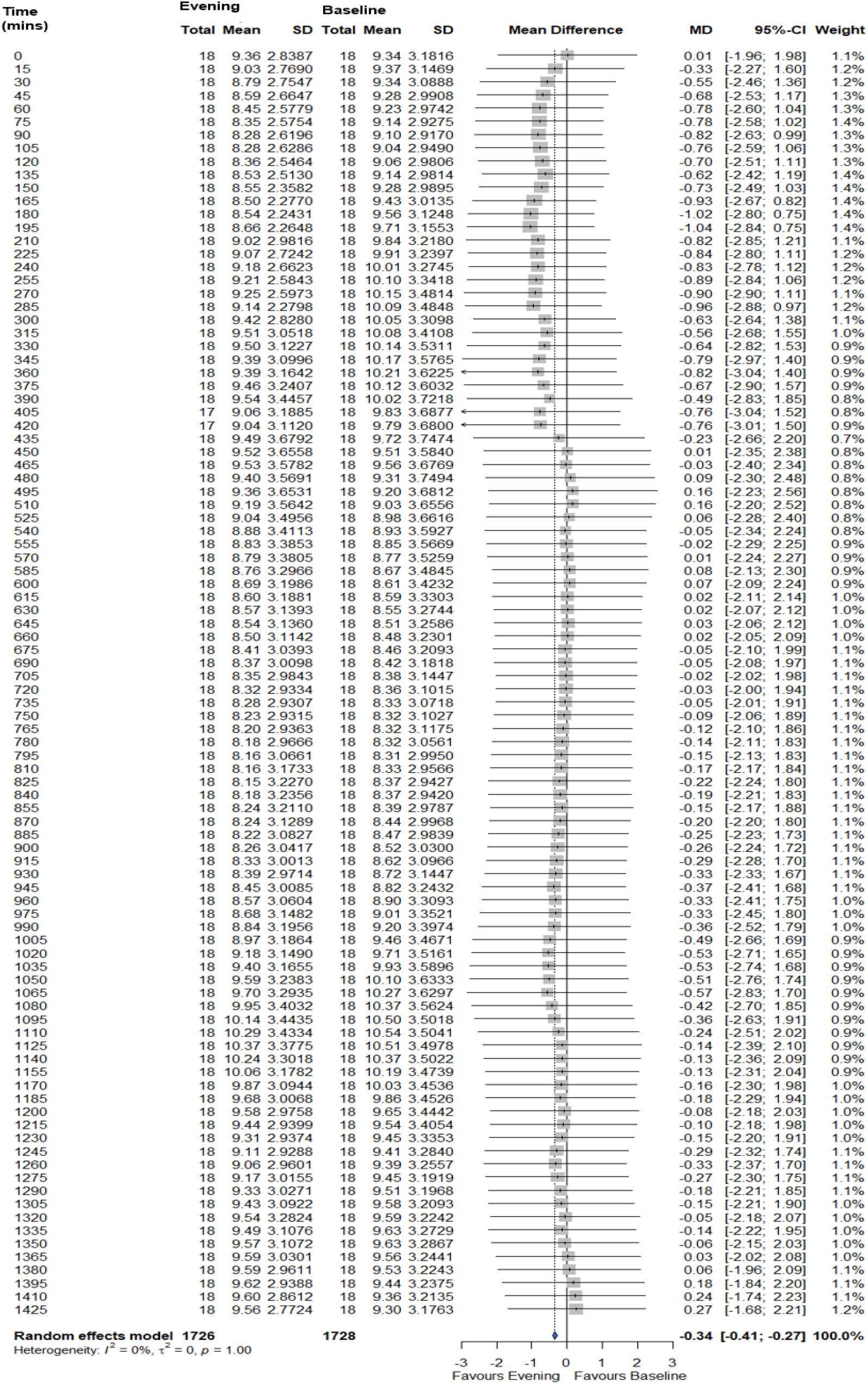
Mean Difference Meta-Analysis Comparing Baseline and Evening Exercise Intervention: 24-hour period is split into 15-minute intervals. Here, baseline is time matched to evening exercise intervention therefore 4pm = 0 mins. Evening exercise period ends at 7pm (180 mins). A negative MD favours evening and positive MD favours baseline. SD (standard Deviation), MD (Mean Difference).

Morning exercise compared to baseline had an overall mean difference in AUC glycaemia (and 95% confidence intervals) of -0.63 (-0.68, -0.58) mmol/L per time, with paired t-tests showing statistical significance, t(94) = 28.18, p<0.001. Evening exercise compared to baseline also showed statistical significance with a paired t-test of t(95) = 2.44, p = 0.016 and a mean difference and 95% confidence interval of -0.34 (-0.41, -0.27) mmol/L per time.The mean difference between baseline and exercise in the evening intervention is lower than the morning intervention. It is also interesting to note that at no point in the day do glucose levels of the morning exercise intervention rise to similar or higher values recorded in baseline intervention. Our data demonstrates that 88% of participants recorded lower daily average glucose with morning exercise compared to evening exercise (57% week 1-2 of morning intervention and 43% week 5-6 of morning intervention). The results also show that some individuals respond more effectively to exercise intervention and metformin intake compared to others.

### Interaction Between Metformin Intake and Exercise

Figure 8 shows the mean value of AUC glucose for those who recorded their metformin intake in both morning and evening exercise interventions. Unpaired t-tests showed there was a significant difference between the AUC values of those who took metformin pre-breakfast rather than post-breakfast during morning exercise intervention (t (11) = 4.188, p=0.002). The same significant difference was not seen between pre and post-breakfast intake of metformin with evening exercise (t (11) = 1.101, p=0.295).

**Figure 8.**
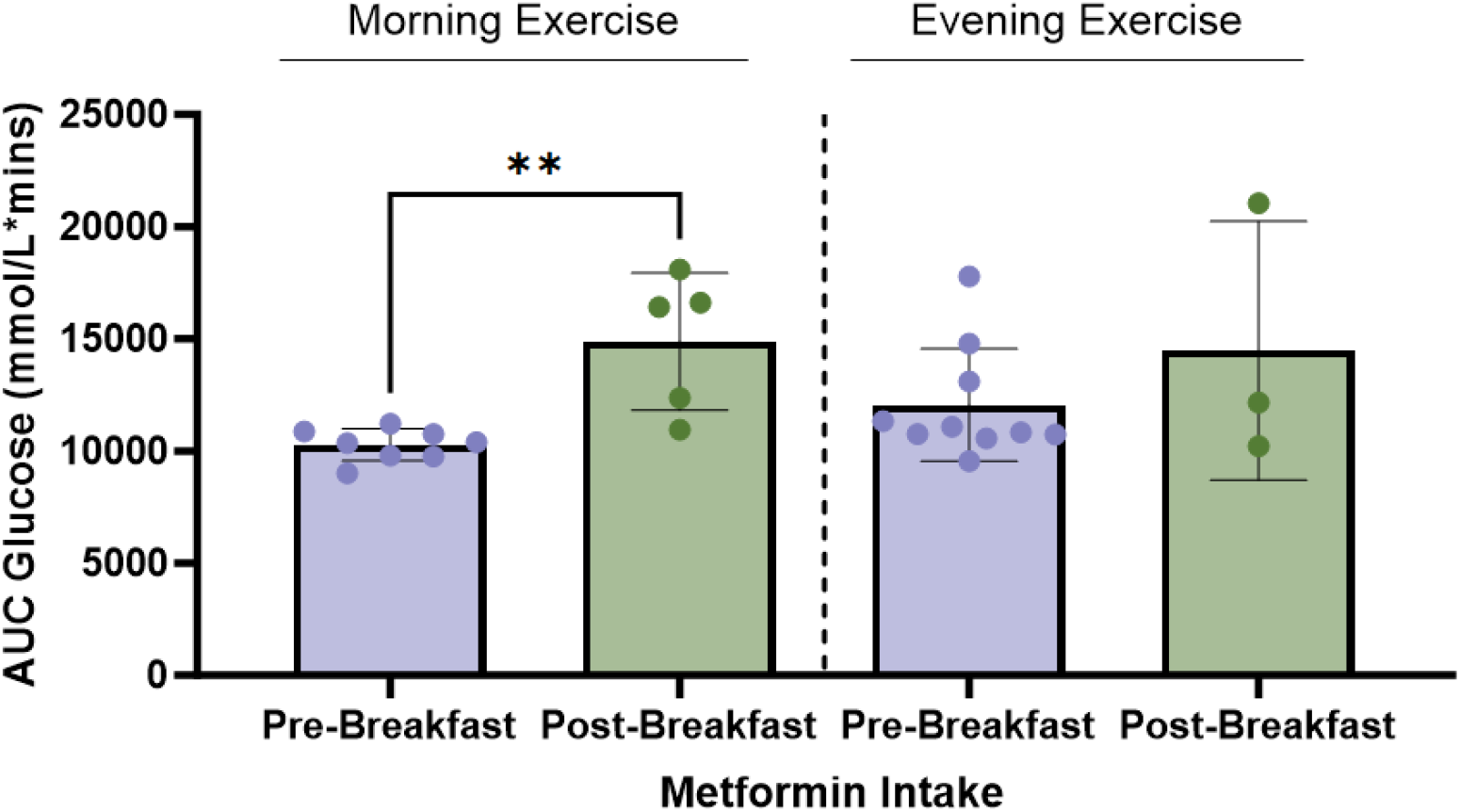
Difference in AUC values between different metformin intake routine during morning and evening exercise: Mean plotted with standard deviation showing the difference in AUC (Area Under Curve) values when metformin intake was recorded pre-breakfast and post-breakfast in both morning and evening exercise interventions. ** denotes significance of P-value < 0.01.

Table 2 records when each participant took metformin dose in each exercise intervention and during baseline.

**Table 2.** Timing of Metformin Intake during each Intervention Period: Table highlights if metformin was taken pre-breakfast or post-breakfast during each exercise intervention for each participant. The grey boxes indicate when metformin intake timings were not specified by the participant.

| Participant | Baseline | Morning Exercise | Evening Exercise |
| --- | --- | --- | --- |
| Participant 1 |  |  |  |
| Participant 2 | Pre-breakfast | Post-breakfast | Pre-breakfast |
| Participant 3 | Post-Breakfast | Post-breakfast | Post-Breakfast |
| Participant 4 |  |  |  |
| Participant 5 | Pre-breakfast | Pre-breakfast | Pre-breakfast |
| Participant 6 | Post-breakfast | Post-breakfast | Post-breakfast |
| Participant 7 | Pre-breakfast | Pre-breakfast | Pre-breakfast |
| Participant 8 |  |  |  |
| Participant 9 |  |  |  |
| Participant 10 |  | Post-Breakfast | Pre-breakfast |
| Participant 11 | Post-breakfast | Post-Breakfast | Pre-breakfast |
| Participant 12 | Pre-breakfast | Pre-breakfast | Pre-breakfast |
| Participant 13 | Pre-breakfast | Pre-breakfast | Post-breakfast |
| Participant 14 | Pre-breakfast | Pre-breakfast | Pre-breakfast |
| Participant 15 | Pre-breakfast | Pre-breakfast | Pre-breakfast |
| Participant 16 | Post-breakfast |  | Post-Breakfast |
| Participant 17 | Pre-breakfast | Pre-breakfast | Pre-breakfast |
| Participant 18 | Pre-breakfast | Pre-breakfast | Pre-breakfast |

56% of participants remained consistent in their timings of metformin intake throughout the study and 22% of participants altered their metformin timings between pre and post-breakfast for morning and evening exercise. 22% did not specify the timing of metformin intake.

Figures 9-10 show the timing of exercise and metformin associated with an individual’s lowest recorded AUC 24-hours glycaemic value. Overall, 67% of the cohort recorded a lower AUC 24-hours glycaemic value in response to exercise, and 33% had a lower AUC 24-hours glycaemic value during the baseline period. Among those with a lower AUC value in response to exercise, 75% had a lower AUC value with morning exercise compared to 25% with evening exercise. 50% of the cohort recorded a lower AUC value with pre-breakfast metformin intake. The combination of pre-breakfast metformin with morning exercise resulted in an optimal glucose response in 5 participants, the most of any combination.

**Figure 9.**
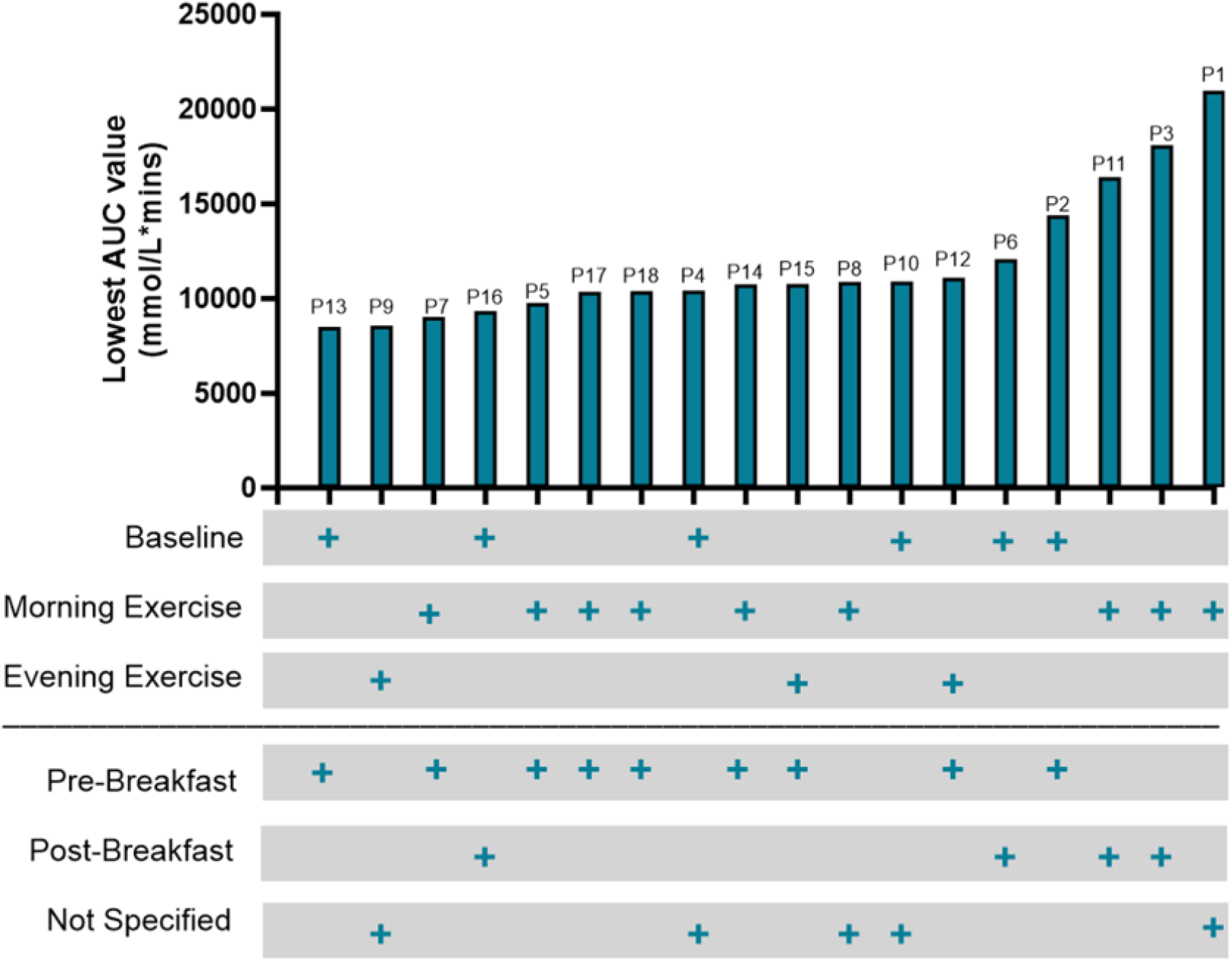
Optimal Exercise timing and metformin intake which gives the lowest AUC value for each participant: Lowest area under the curve (AUC) value obtained from table 2 is matched to the intervention period and recorded metformin intake obtained from table 3.

**Figure 10.**
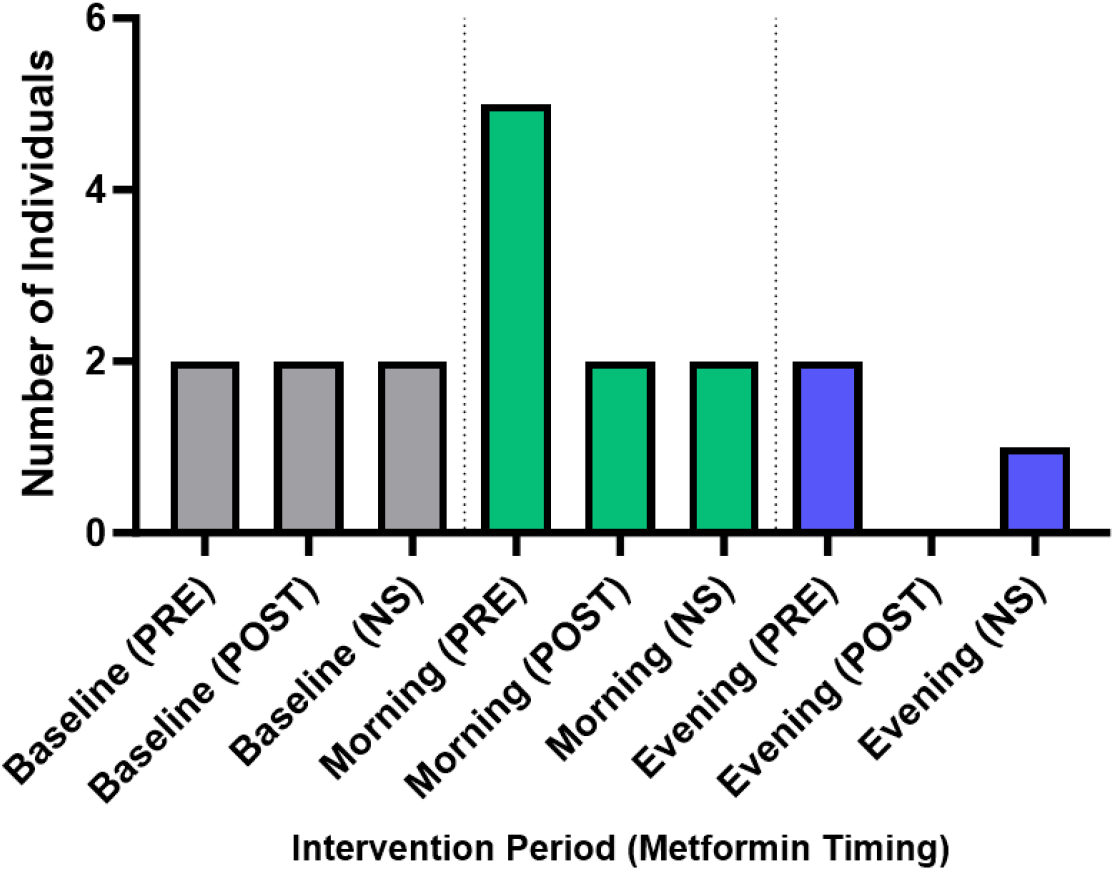
Number of Individuals with optimal glucose response in each intervention period with metformin timing: Summary of results obtained from figure 9, highlighting the number of individuals with optimal responses to each exercise treatment and metformin timing. Pre-Breakfast (PRE), Post-Breakfast (POST), Not Specified (NS). Graph made on GraphPad Prism.

## Discussion

Researchers have typically employed observational, cross-sectional, and parallel-group randomized trials to study circadian biology and disease management, but these approaches may not reliably identify the most effective treatments for specific individuals or subgroups. We have used a n-of-1 approach to analyse a previously published circadian cross-over trial performed in people with type 2 diabetes. This 16-week randomised crossover study was designed to investigate glycaemic outcomes in response to moderate intensity exercise at different times of the day in people with Type 2 Diabetes also being prescribed metformin monotherapy. Secondary outcomes included total physical activity, medication timing, diet and sleep. The trial was designed to determine if it is possible to optimise timing of concomitant metformin and exercise in people with type 2 diabetes. An n-of-1 approach has allowed us to reveal heterogeneity within this cohort and identify individuals that appear to respond more beneficially to one or both arms of the exercise intervention. This approach has also allowed identification of individuals that responded to concomitant timings of metformin intake in relation to meal timing.

The findings indicate that morning exercise is more effective than evening exercise in lowering blood glucose levels over a 24-hour period post-exercise in people with type 2 diabetes who are being prescribed metformin monotherapy. Both morning and evening exercise reduced overall mean blood glucose levels, but morning exercise had a significantly larger effect. During the first two weeks, morning exercise had a greater impact on reducing glucose levels than in weeks five to six. Most participants experienced lower daily average glucose levels with morning exercise compared to evening exercise. Individual responses varied, with some participants consistently showing large reductions in glucose levels with both morning and evening exercise, while others only showed this response to one. Improvements to glucose levels in response to exercise was not apparent for some participants whose glucose levels were optimal at baseline. Participants self-reported when they consumed metformin which allowed us to analyse the most optimal timing for metformin dosage in relation to exercise and dietary intake timing. Most participants remained consistent in their metformin intake timings throughout the trial. Similar to our previous findings (Carrillo *et al*., 2024), taking metformin before breakfast combined with morning exercise significantly lowered AUC glucose values, an effect not seen with evening exercise. When analysing individuals across the study overall, the optimal combination for the lowest AUC values appeared to be the combination of morning exercise and pre-breakfast metformin intake, which was the most effective combination for 5 participants, the most of any combination. A significant portion of the cohort responded best to morning exercise, while a smaller percentage displayed a more optimal response with evening exercise or during the baseline period. Our data also reveals that regardless of Zeit-time normalisation i.e. whether glucose time-courses are analysed by time-of-day, or with exercise as the start of a 24 time-course (Zeit-time normalised), morning exercise lowers glycaemia more efficaciously than evening exercise across this cohort. Zeit-time normalisation analysis removes confounding factors such as lag-time of beneficial effects of exercise upon glycaemic regulation. Previous studies have not employed this normalisation technique when assessing time-of-day exercise outcomes (Savikj *et al*., 2019; Mancilla *et al*., 2021; Moholdt *et al*., 2021; van der Velde *et al*., 2023; Qian *et al*., 2023). It will be beneficial for future work that assesses diurnal exercise outcomes to analyse data using a standard time-of-day comparison, in addition to a Zeit-time normalised analysis to mitigate against confounding factors.

A limitation of the study is that it was not originally designed in a n-of-1 format. However, due to the nature of the crossover design and the inclusion of continuous data monitoring in the form of continuous glucose monitors, the study is suitable for post-hoc analysis of this kind. Remote monitoring is common practice in n-of-1 studies however, the use of wearable technology increases participant burden. Individual exercise intervention adherence may play a role in the differential responses found in this study. Adherence was monitored in real-time and participant data was excluded from the final data analysis if they met any of the following criteria: missing >4 exercise windows in less than 2 weeks, no glucose readings for >4 days, or if participants had a heart rate discrepancy of >15% during exercise. Despite these adherence criteria, we cannot rule out the possibility that individual differences in adherence influence our findings.

The biology that underlies circadian heterogeneity in metabolism is not fully understood. The molecular clock influences various aspects of circadian metabolism at the cellular level and helps regulate metabolite uptake by peripheral tissues, including glucose (Gabriel & Zierath, 2021). Exercise has the ability to reset the molecular clock in peripheral tissues (Wolff & Esser, 2012), and molecular clock mutations are found in individuals with irregular circadian rhythms, such as those with familial delayed sleep phase disorder (Patke *et al*., 2017). Thus, disturbances to the molecular clock caused by morning or evening exercise may partly explain the individual variability in glycaemic outcomes observed in this study. Skeletal muscle is responsible for the majority of postprandial glucose uptake and plays a major role in regulating glycaemia (DeFronzo & Tripathy, 2009). It has been observed that people with type 2 diabetes have less robust intrinsic skeletal muscle circadian rhythms at the molecular clock and mitochondrial levels (Gabriel *et al*., 2021). These ablated circadian rhythms may lead to increased heterogeneity in diurnal exercise responses in this group, since differences in diurnal exercise glycaemic responses appear to be of a smaller magnitude in people with normal glucose tolerance (Tanaka *et al*., 2021).

In this study we have investigated the heterogeneity in response to the timing of exercise and metformin intake using an n-of-1 approach within a randomised crossover trial. This analysis technique may allow for individualised chrono-medicine strategies. We have shown that morning exercise combined with pre-breakfast metformin intake is the most effective strategy for lowering blood glucose levels in the greatest number of participants with type 2 diabetes in both a real-time and Zeit-time normalised analysis. However, individual response heterogeneity may benefit from a personalised strategy via chrono-medicine. Furthermore, a holistic approach to chrono-medicine is necessary for optimal health outcomes, including assessment of the diurnal environment in which an individual exists.

## Data Availability

Data availability
Raw data underlying the original manuscript have been deposited publicly https://doi.org/10.6084/m9.figshare.24081417.
Code used to re-analyse these data have been publicly deposited at https://github.com/kmlyons13/N-of-1-Analysis-on-Circadian-Data.

https://doi.org/10.6084/m9.figshare.24081417.

https://github.com/kmlyons13/N-of-1-Analysis-on-Circadian-Data.

## Acknowledgements

N/A

## Author contributions

K.L., B.J.P.C., and B.M.G contributed to the conception or design of the work. K.L., B.J.P.C., L.T.D., O.P.I, and B.M.G contributed to acquisition, analysis or interpretation of data. All authors contributed to drafting the work or revising it critically for important intellectual content. All authors have read and approved the final version of this manuscript and agree to be accountable for all aspects of the work in ensuring that questions related to the accuracy or integrity of any part of the work are appropriately investigated and resolved. All persons designated as authors qualify for authorship, and all those who qualify for authorship are listed.

## Data availability

Raw data underlying the original manuscript have been deposited publicly https://doi.org/10.6084/m9.figshare.24081417.

Code used to re-analyse these data have been publicly deposited at https://github.com/kmlyons13/N-of-1-Analysis-on-Circadian-Data.

